# Polygenic prediction of treatment efficacy with causal transfer learning

**DOI:** 10.1101/2025.10.15.25338051

**Authors:** Jiacheng Miao, Jin Mu, Xiaoyu Yang, Jason M. Fletcher, Lauren L. Schmitz, Qiongshi Lu

## Abstract

Therapeutic interventions often exhibit heterogeneous treatment effects (HTE) across individuals. A central goal of precision medicine is to enable personalized treatment recommendations based on patients’ measurable characteristics. Identifying factors that explain HTE is therefore essential. However, detecting HTE remains challenging due to limited sample size in randomized controlled trials (RCTs), often-missing baseline information, and suboptimal statistical methods with limited power. Here, we introduce a principled statistical framework named M-Learner to identify genetically-driven HTE. This approach leverages genetic variation involved in diverse biological pathways influencing drug response, integrates insights from two decades of complex trait genetics, and employs causal transfer learning applicable to both individual-level data and summary statistics. Applying M-Learner to multiple RCTs, we found low bone mineral density as a key determinant of secukinumab efficacy in ankylosing spondylitis, and identified smoker subpopulations adversely affected by a bronchodilator treatment. Our findings demonstrate the utility of genetic variation in HTE inference and make important advances toward the promise of precision medicine.

## 1 Introduction

Heterogeneous treatment effect (HTE) refers to the variation in the effect of a treatment across individuals or subpopulations [1]. Pinning down the source of HTE will have profound implications in precision medicine, which seeks to customize healthcare based on individual characteristics [2, 3]. With a full understanding of how an individual’s (pre-treatment) characteristics affect treatment response, healthcare providers can optimize the treatment strategy for each patient based on these characteristics, potentially enhancing efficacy and minimizing adverse effects [4].

For example, secukinumab (Cosentyx) is an anti-interleukin-17 (IL-17) monoclonal antibody widely used for treating inflammatory diseases [5]. It has demonstrated effectiveness across several conditions, but is also known to exhibit considerably variable therapeutic outcomes, with response rates ranging from 81.6% for psoriasis [6], 62.6% for psoriatic arthritis [7], 60.5% for ankylosing spondylitis [8], to only 30.7% for rheumatoid arthritis [9]. These moderate treatment response rates demonstrate a critical need for more individually-tailored therapeutic approaches. Efforts have been made to identify factors that can explain the variation in secukinumab treatment response, but have had limited success [10].

This reflects a broader challenge in precision medicine research: detecting HTE is inherently difficult. Randomized controlled trials (RCTs) often have limited sample size and are primarily designed to estimate average treatment effects (ATE) rather than HTE. In addition, variables driving the heterogeneity in treatment response are unknown prior to the trial and can be highdimensional. Therefore, these variables are often missing in RCT data, even though some trials may be sufficiently powered to identify HTE.

In this paper, we introduce two key advances to the current practice of HTE inference. First, we use genome-wide genetic variation to predict HTE—a paradigm we term “polygenic efficacy”. A main reason why we envision genetics playing a key role in HTE inference is that germline DNA genotypes are fixed at conception. Therefore, post-trial genetic measures are identical to the baseline measures since the treatment will not change DNA genotypes. This means that, with proper consent in place, it is possible to retrospectively sequence RCT participants, creating highdimensional “baseline” genetic measures that are known to associate with nearly every human trait, characteristic, and behavior, which can inform on HTE.

However, genetic data are high-dimensional, with each genetic variant explaining a tiny fraction of variation of most human traits [11]. Therefore, such information needs to be leveraged in conjunction with advanced new methods that can handle infinitesimal genetic effects. Few studies have explored using genetic information to infer HTE [12], and existing studies have adopted either subgroup analysis or risk score analysis [13]. For example, a recent study investigated whether genetic variables modify the therapeutic effects of secukinumab in four inflammatory diseases [10]. Their subgroup analysis stratifies the study sample based on each variant’s genotype and compares treatment effects across genotype groups [4], which is underpowered due to the small trial sample, modest variant effect size, and high burden of multiple testing. The risk score analysis aggregates genome-wide genetic variation into polygenic risk scores (PRS) [14–16], which predict the risk of developing immune diseases rather than treatment response, and tested whether PRS modifies treatment efficacy [10, 17–19]. This strategy also has major limitations since the PRS developed without using any information on the treatment is not guaranteed to correlate with treatment effectiveness. Indeed, both analyses produced null results. The second key advance we introduce in our study is to develop a principled statistical approach, named MLearner, to quantify genetic influences on HTE. It is a causal transfer learning framework for HTE estimation that accommodates both individual-level genetic data and summary-level association statistics. We demonstrate its utility using RCTs on anti-IL17 monoclonal antibodies for inflammatory diseases and an RCT on a bronchodilator treatment for better lung function in smokers. These new approaches align with the growing trend towards more personalized healthcare solutions, promising to enhance patient care by tailoring treatment options to the unique genetic makeup of every individual.

## 2 Results

### Statistical formulation of polygenic efficacy

We first introduce the concept of polygenic efficacy, defined as the aggregated contribution of genome-wide genetic variation to treatment response. To formulate this concept, we adopt a polygenic genotype-by-treatment interaction (G×T) model motivated from recent advances in the literature on gene-environment interplay [20]. We also present a more general, nonlinear extension of this framework in the Methods section:

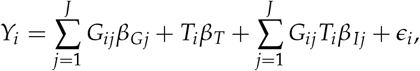

where *Y*_*i*_ is the treatment outcome (which can be a measure of treatment efficacy or adverse effect, depending on the application) for individual *i, G*_*ij*_ is the standardized genotype at variant *j* (mean 0 and variance 1), *T*_*i*_ is the standardized treatment indicator with treatment probability *p, β*_*T*_ is the treatment effect, *β*_*Gj*_ and *β*_*Ij*_ denote the additive and interaction effects for variant *j*, and *ϵ*_*i*_ is the error term. We define the polygenic efficacy score (PES) for individual i as the weighted sum of allele counts with weight values given by their G×T effects.

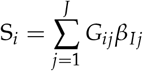

Conditional average treatment effect (CATE) is a commonly used measure of HTE. It is defined as the expected outcome difference between treatment and control groups given a set of covariates [21]. In our study, the covariates are the allele counts of millions of single-nucleotide polymorphisms (SNPs) in the genome (i.e., *G*_*ij*_, *j* = 1, …, *J*). Under the polygenic G×T model, it can be shown that CATE for individual i is a linear function of the PES (Methods), linking genome-wide GxT interactions directly to HTE:

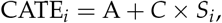

where A is the ATE and C is a positive constant related to the probability *p* of being assigned to the treatment group of the trial. Therefore, PES (i.e., S_*i*_) explains all the variability in treatment response that is attributable to genetic variation. If larger *Y*_*i*_ indicates better outcomes, individuals with higher *PES* values are expected to experience greater treatment benefit. We note that the concept of polygenic efficacy naturally extends to polygenic safety, which captures the aggregated genetic contribution to the risk of developing adverse events or treatment-related harms.

From these derivations, it follows that estimating genetically-driven HTE is equivalent to estimating the PES. However, this task can be challenging in an RCT because of limited sample size and the typically small GxT effect at the SNP level. In fact, as we will illustrate later, directly estimating SNP weights for a PES (i.e., *β*_*Ij*_) from RCT data is nearly impossible. To address this challenge, we introduce M-Learner, a causal transfer learning framework that leverages the power of PRS models pre-trained from well-powered GWAS.

### An overview of M-Learner

M-Learner is built on two key ideas: (1) it models polygenic efficacy as a (possibly nonlinear) function of high-dimensional PRS variables (Figure 1A), and (2) it employs a causal transfer learning framework that fine-tunes pre-trained PRS models in RCT data to predict treatment response. This design moves beyond conventional strategies that rely on a handful of candidate variants or a single PRS for the trial outcome, which do not fully capture the complex role of genetic background in HTE. By compressing millions of SNPs into PRS variables, M-Learner leverages external information from many exceptionally-powered GWAS and alleviates the computational challenge in modeling high-dimensional genetic data. Later, we will demonstrate that these advances address the limited statistical power inherent to small RCTs.

**Figure 1:**
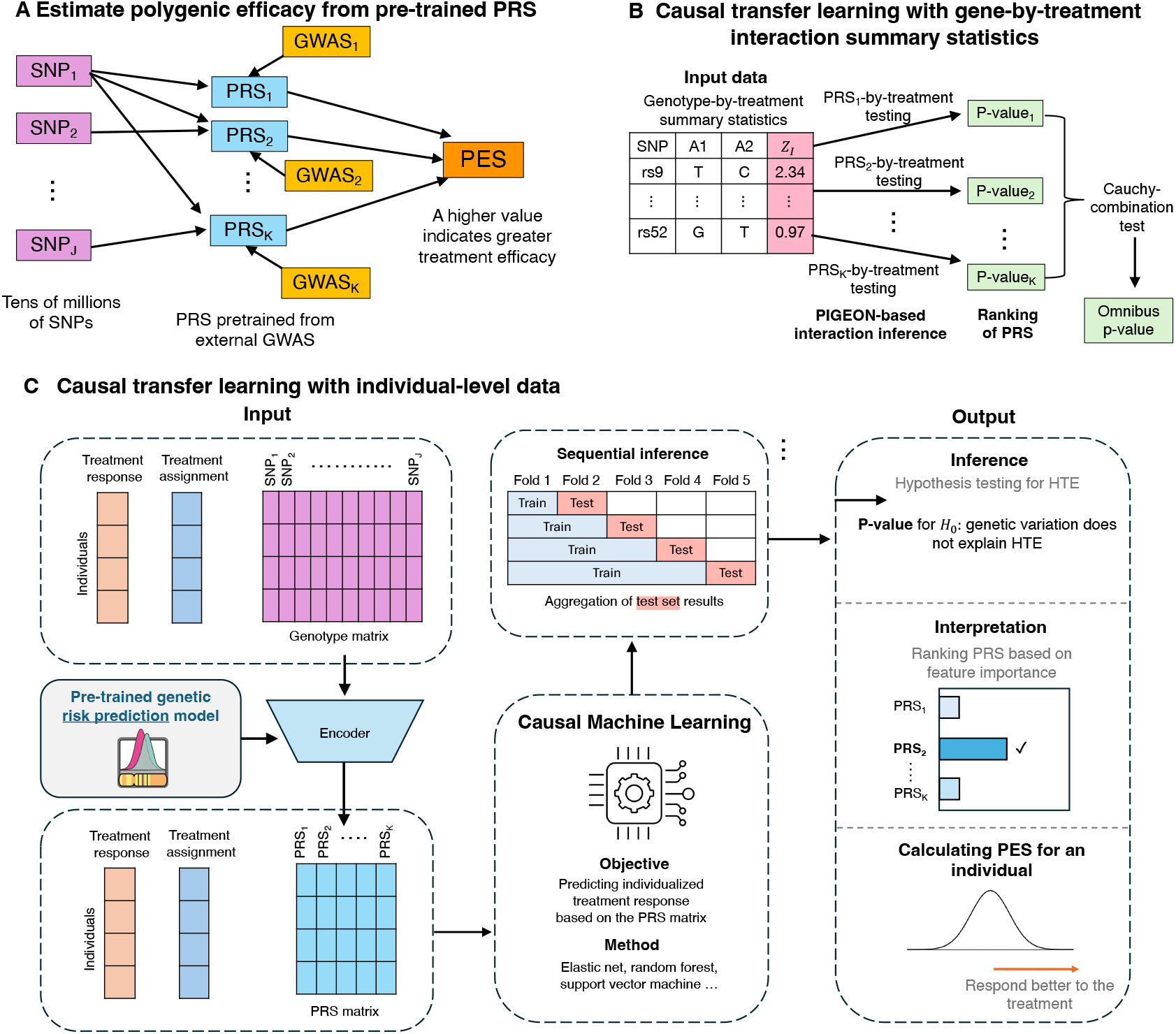
M-Learner framework for modeling polygenic efficacy and HTE. **(A) Genetic representation of polygenic efficacy.** Instead of relying on candidate variants or a single PRS for the trial outcome, M-Learner uses many PRS derived from external GWAS to capture the complex genetic mechanisms underlying HTE. **(B) M-Learner based on summary statistics (M-Learner-S)**. Using GxT GWIS summary statistics, M-Learner tests many PRS’ interactions with the treatment using PIGEON and integrates them with the Cauchy combination test. The output includes a p-value for testing the HTE and a ranked list of PRS that are informative on the HTE. **(C) M-Learner based on individual-level data (M-Learner-I)**. M-Learner applies causal transfer learning to fine-tune pretrained PRS models for individualized treatment prediction. Sequential inference ensures valid statistical testing. The outputs include a P-value for detecting HTE, a ranked list of PRS contributing to HTE, and individualized predictions of treatment response.

We introduce two versions of M-Learner to accommodate different types of input data: M-Learner-S, which uses GxT summary statistics from genome-wide interaction studies (GWIS) in settings with only privacy-preserving summary-level data available (Figure 1B), and M-Learner-I, which leverages individual-level genotype and treatment data for more sophisticated investigation (Figure 1C). Briefly, M-Learner-S first employs a polygenic interaction inference approach to conduct a hypothesis-free scan of PRS-by-treatment interactions from summary statistics [20]. The resulting interaction p-values can identify PRS that are informative on the HTE. Results across multiple PRS are then combined using the Cauchy combination test [22] to yield a global P-value. Alternatively, M-Learner-I first compresses millions of SNPs into a matrix of pre-trained PRS models [23, 24]. These PRS, together with treatment assignments and outcomes in the RCT, are then used to fit sophisticated machine learning models for predicting treatment efficacy [25, 26]. We use sequential inference to ensure valid statistical testing while avoiding overfitting [27, 28]. This method produces a global P-value for genetic modification of treatment response, identifies informative PRS through feature importance analysis, and calculates PES that predict how well each patient is expected to respond to the treatment which can be used to design personalized treatment strategies [29].

### Scaling laws for HTE estimation with genetic information

Next, we derive power scaling laws for estimating HTE using genetic variation, focusing on the PES×T interaction coefficient. Here, PES is first estimated through machine learning and then included in the regression to assess its interaction with the treatment, using an independent RCT subsample for evaluation. Intuitively, when the estimated PES×T coefficient is close to zero, there is little statistical power to detect HTE; when it approaches one, the predictive model is well calibrated. The scaling laws provide theoretical guidance on the feasibility of detecting polygenic efficacy in RCTs and highlight the need to leverage pre-trained PRS models for reliable inference.

We compare three strategies for estimating PES: a within-sample GWIS approach, a single-outcome PRS approach, and a multi-PRS ensemble approach (detailed derivations are presented in the Methods section). The within-sample GWIS approach estimates SNP-by-treatment interaction coefficients directly from the RCT samples and uses them as SNP weights for the PES; naturally, limited sample size in the RCT makes these estimates extremely noisy, yielding lower power. The single-outcome PRS approach uses a pre-trained PRS for the trial outcome to assess the interaction with the treatment; its performance depends on the GWAS sample size which determines measurement error in the PRS, as well as the correlation between this PRS and the true PES (which is also equivalent to the correlation between *β*_*Gj*_ and *β*_*Ij*_). Finally, the multi-PRS ensemble approach leverages a high-dimensional PRS matrix pre-trained from many GWAS and fine-tuned in the RCT sample, to predict treatment response. This strategy effectively combines multiple PRS that are informative on the PES and provides improved power to detect genetically-driven HTE.

Figure 2 illustrates these power scaling laws under realistic parameter settings: an RCT training and testing sample size of 2,500, a GWAS sample size of 400,000 for PRS training, a heritability of 0.3 for the trial outcome, and an effective number of 60,000 SNPs. We also fix the type-I error rate at 0.05 in these curves. Panel A shows that without transfer learning (i.e., the within-sample GWIS approach), statistical power depends entirely on the proportion of treatment response variance explained by G×T. Because we expect this parameter to be small (around 0–5% in gene–environment interaction studies), statistical power of this approach is negligible. Panel B fixes the true G×T-explained variance at 3% and examines the effect of transfer learning. Here, power rises steeply as the estimated PES approaches its true value (quantified by the correlation between true and estimated PES), reaching 100% once the correlation reaches moderate levels. If a single PRS weakly correlated with the true PES is used, the analysis will have little power. In contrast, a multi-PRS ensemble approach aggregates information across multiple PRS to obtain a better estimate of the PES and, consequently, achieve higher power. In practice, M-Learner achieves this through an ensemble of PRS pre-trained from external GWAS, enabling detection of genetically-driven HTE even when the G×T signal is weak.

**Figure 2:**
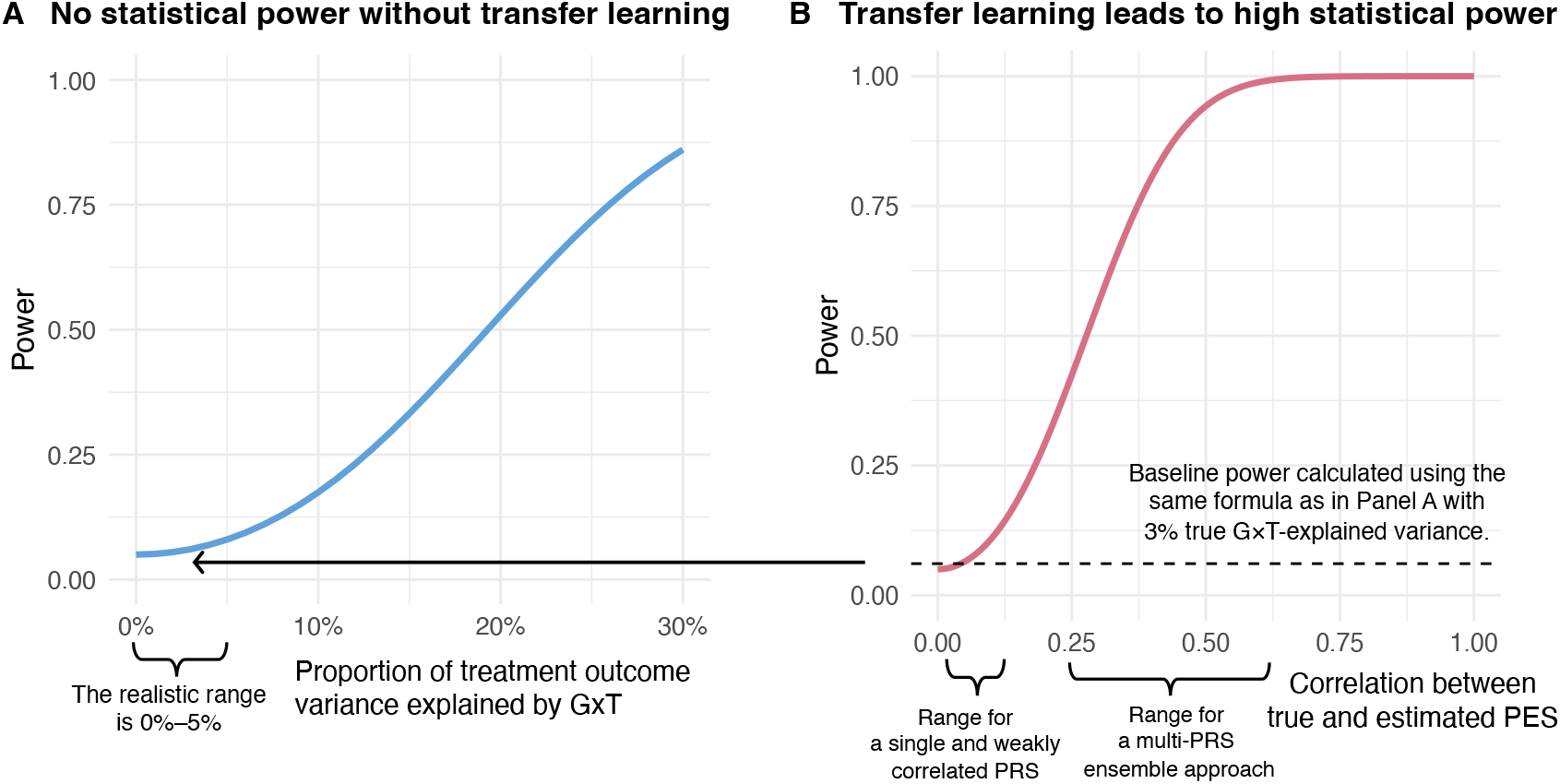
Power scaling laws for detecting genetically-driven HTE. **(A)** For analyses without transfer learning, the graph plots statistical power (y-axis) against the proportion of treatment response variance explained by gene-by-treatment interaction (x-axis). At the realistic range of this value (i.e., 0–5%), statistical power for detecting HTE is negligible. **(B)** With transfer learning, the graph plots power (y-axis) against the correlation between the true and estimated PES (x-axis). When the correlation reaches 0.5, we achieve >90% power. M-Learner achieves high power by aggregating information from a large number of pre-trained PRS.

### Simulation studies

We performed extensive simulations using genotype data from the UK Biobank [30] to compare M-Learner with alternative methods. These simulations were designed to reflect realistic genetic architectures and treatment response models (Methods) [31]. We assessed each method’s ability to detect HTE, reporting statistical power and type I error rates.

Specifically, we simulated a quantitative treatment outcome under a polygenic G×T interaction model with 200,000 SNPs. SNP effect sizes were drawn jointly from a multivariate normal distribution. The G×T term was specified in both linear and nonlinear forms. RCT sample sizes of 500, 1,000, and 2,000 individuals were examined. Each simulation scenario was repeated 500 times. Power was defined as the proportion of replicates rejecting the null hypothesis of no HTE at a significance level of 0.05, while type I error was evaluated under settings without true HTE. We considered four alternative methods: (i) Single-outcome PRS, which uses the trial outcome PRS to approximate PES; (ii) Within-sample GWIS, which constructs the PES directly from the SNP-by-treatment interaction coefficients; (iii) PRSPGx [32], which constructs the PES as a weighted sum of the outcome PRS and GWIS, where the weights are selected to maximize the prediction of treatment response in hold-out samples. (iv) M-Learner (SNP): which employs M-Learner to estimate CATE directly from SNP genotype data without transfer learning. Here, methods (i) and (ii) are two approaches we have compared with in the scaling law discussion, showcasing the performance of using either a single PRS on the outcome or a (noisy) PES estimated from RCT as the genetic modifier of treatment effect. Method (iii) is a combination of methods (i) and (ii) without using high-dimensional PRS information or machine learning. Method (iv) explores the performance of M-Learner without external PRS models. This is a comprehensive list of available strategies for estimating HTE from genetic data, and serves as an important benchmark for M-Learner’s performance.

All methods maintained well-calibrated type I error rates in both linear and nonlinear settings (Figure 3A-B), but M-Learner achieved substantially higher statistical power than other approaches in all settings. Among the alternatives, the single-outcome PRS approach demonstrated modest power in the linear setting but failed to capture nonlinear effects. The other three methods showed low power in all scenarios. Together, these findings demonstrate M-Learner’s ability to identify genetically-driven HTE in small samples that are comparable to real-world RCTs, especially under a nonlinear mapping from PRS to treatment effectiveness, where other approaches fail. These results also clearly suggest that the strategy to incorporate many PRS from external GWAS is a key methodological advance that improves HTE inference.

**Figure 3:**
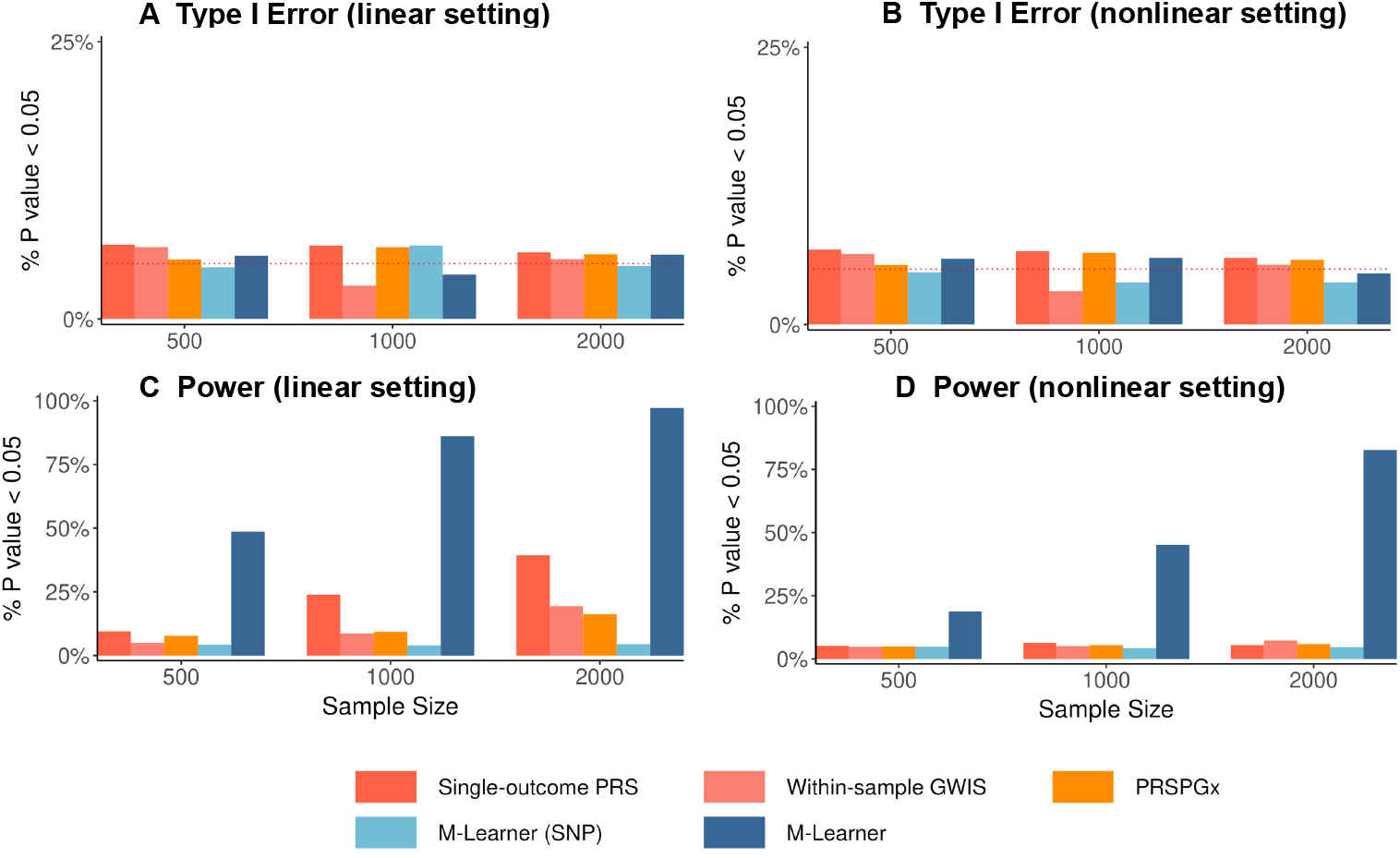
Simulation results comparing M-Learner with alternative methods for detecting HTE. Panels A and B show that all methods maintain well-calibrated type I error around the nominal 0.05 level under both linear (A) and nonlinear (B) settings. Panels C and D demonstrate that M-Learner achieves substantially higher power than alternative approaches across increasing sample sizes, with consistent gains under both linear (C) and nonlinear (D) settings. Competing methods exhibit limited power, especially in nonlinear scenarios, underscoring the advantage of M-Learner in detecting genetically driven heterogeneous treatment effects.

### Genetic factors explain the HTE of secukinumab in ankylosing spondylitis

Next, we revisit the heterogeneous response to the anti-IL17 monoclonal antibody secukinumab in inflammatory diseases. A recent study failed to identify genetically-driven HTE, but released GWIS summary statistics for SNP-by-treatment interactions on four diseases (i.e., psoriasis, psoriatic arthritis, rheumatoid arthritis, and ankylosing spondylitis) from 19 RCTs (total N_treated_=4,063, N_control_=1,151; Supplementary Table 1) [10]. We applied M-Learner-S to investigate whether the HTE of secukinumab can be explained by genetic variation. Following previous work, we investigated four continuous measures of disease activity as the primary outcomes, including the disease activity score 28 with C-reactive protein (DAS28-CRP) for psoriatic arthritis and rheumatoid arthritis, the ankylosing spondylitis disease activity score with C-reactive protein (ASDAS-CRP), and the psoriasis area and severity index (PASI) score [10]. We used 70 external GWAS for a wide range of complex traits as inputs to M-Learner-S (Supplementary Table 2).

The previous study used conventional one-SNP-at-a-time subgroup (GWIS) analysis and risk score analysis based on the PRS of 15 immune diseases and found null results [10]. Using their GWIS summary statistics, we reproduced these null PRS×treatment interaction results within the M-Learner framework (Supplementary Figure 1). Next, we employed M-Learner to identify genetically-driven HTE. We identified significant HTE driven by genetic variation in the primary outcome for ankylosing spondylitis, i.e., ASDAS-CRP, after multiple testing correction (omnibus P = 0.01) (Figure 4A). To follow up on this primary finding, we employed M-learner to examine two secondary outcomes for ankylosing spondylitis, i.e., C-reactive protein (CRP) and erythrocyte sedimentation rate (ESR). We found highly significant HTE on CRP and suggestive results on ESR (P = 0.0001 for CRP and P = 0.086 for ESR). We did not identify significant HTE for psoriasis, psoriatic arthritis, or rheumatoid arthritis.

**Figure 4:**
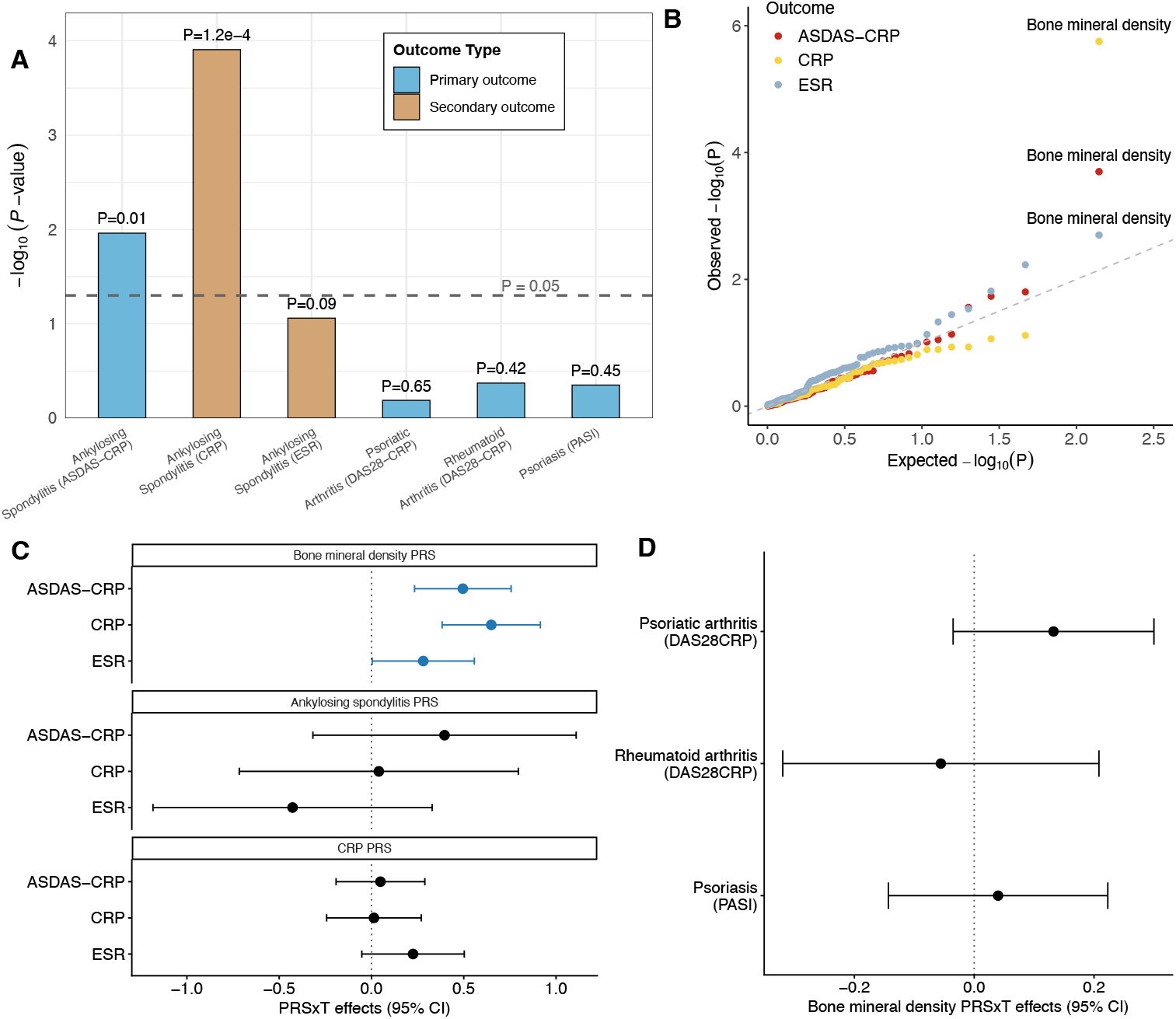
M-Learner identifies the PRS of BMD as the top predictor of secukinumab HTE in ankylosing spondylitis. **(A)** M-Learner detects significant genetically driven HTE for ankylosing spondylitis (P = 0.01 for ASDAS-CRP and P = 1.2e-4 for CRP) (Supplementary Table 3), but not for other three diseases. **(B)** QQ plot for the PRSxT interaction p-values on three outcomes of ankylosing spondylitis. Each dot represents a PRS. **(C)** Individuals genetically predisposed to lower BMD show greater benefit from anti-IL17 treatment for ankylosing spondylitis. ASDAS-CRP: Ankylosing spondylitis disease activity score with CRP; CRP: C-reactive protein levels; ESR: erythrocyte sedimentation rate. Lower value of those treatment outcomes indicates better treatment response. **(D)** BMD PRS does not modify the treatment efficacy for other three diseases.

Next, we sought to identify specific genetic traits that can explain the observed HTE in ankylosing spondylitis. The highest-ranked PRS was bone mineral density (BMD, Figure 4B). This analysis suggests that ankylosing spondylitis patients who had lower PRS of BMD showed better outcomes after treatment, quantified by a greater reduction of ASDAS-CRP (estimated PRSxT coefficient = 0.495, P = 0.0002; Figure 4B-C and Supplementary Table 3). This interaction was also found in the two secondary outcomes CRP (estimated PRSxT coefficient = 0.649, P = 1.8e-6; Figure 4B-C and Supplementary Table 4) and ESR (estimated PRSxT coefficient = 0.281, P = 0.048; Figure 4B-C and Supplementary Table 5). Interestingly, we note that the HTE of secukinumab on ankylosing spondylitis was not explained by the genetic predisposition to ankylosing spondylitis itself or by the trial outcome CRP. PRS of ankylosing spondylitis or CRP did not show significant interactions with the treatment on primary or secondary outcomes (Figure 4C and Supplementary Tables 3-5). The interaction between BMD PRS and treatment on ASDAS-CRP remained significant after conditioning on the PRS of ankylosing spondylitis (estimated PRSxT coefficient = 0.477, P = 0.0007) and the PRS of CRP (estimated PRSxT coefficient = 0.903, P = 5.2e-5). This clearly demonstrates an important point we have made throughout the paper––risk score analysis based on a single PRS relies on a strong assumption that the PRS included in the analysis is correlated with the PES underlying HTE. In this example, although it seemed natural to use the PRS of ankylosing spondylitis (the disease of interest) or CRP (the trial outcome) as the risk score, they do not explain the HTE. The multi-PRS strategy implemented in M-Learner was able to leverage information from many other traits, and in this case, identified a specific interactor BMD that significantly modifies the effect of secukinumab on ankylosing spondylitis. We also note that this is a finding specific to ankylosing spondylitis. BMD did not modify the treatment response in the other three inflammatory diseases in our analysis (Figure 4D).

The findings suggest that the higher treatment benefits to anti-IL17 therapy in patients genetically predisposed to lower BMD may be explained by the central role of IL17 in osteoimmunology. IL17 amplifies bone resorption by upregulating RANKL expression in osteoblasts and stromal cells, inducing downstream cytokines such as TNF, IL-1*β*, and IL-6, and reducing osteoprotegerin (OPG), thus increasing the RANKL:OPG ratio and promoting osteoclastogenesis [33]. Concurrently, IL17 perturbs osteoblast regulation by suppressing the Wnt inhibitors DKK1 and sclerostin, which enhances osteogenic differentiation but in an aberrant, inflammatory context that favors pathological syndesmophyte formation rather than healthy bone accrual [34]. Patients with genetic predisposition to low BMD—characterized by variants that increase RANKL or decrease OPG, and by heightened activity of Wnt inhibitors—are therefore particularly vulnerable to IL17–driven imbalances in bone remodeling [35]. In such individuals, IL17 inhibition not only suppresses the inflammatory cytokine network that fuels osteoclast activation but also restores normal regulation of osteoblast differentiation, leading to both improved systemic bone density and attenuation of pathological new bone formation. This mechanistic framework provides a plausible explanation for the observed HTE, wherein genetically low-BMD patients derive disproportionate clinical and skeletal benefit from anti-IL17 therapy.

### Genetically-derived HTE of the bronchodilator treatment in the Lung Health Study

The Lung Health Study was an RCT conducted across multiple centers in the United States and Canada to evaluate whether a smoking intervention and the long-term use of an inhaled bronchodilator could slow the progressive decline of lung function in smokers with early-stage chronic obstructive pulmonary disease [36]. The trial concluded that smoking cessation substantially reduced the age-related loss of lung function, whereas the bronchodilator did not slow the decline of forced expiratory volume in one second (FEV1), suggesting the limited effectiveness of pharmacologic therapy compared to behavioral intervention [37].

We applied M-Learner to the Lung Health Study to revisit its null finding on the bronchodilator. We included 2,441 participants with both genotype and outcome data, comprising 1,249 in the treatment group (smoking intervention plus bronchodilator) and 1,192 in the control group (smoking intervention plus placebo). The outcome was defined as the cumulative change in postbronchodilator FEV1 over five years. We employed SNP-level subgroup analysis (i.e., GWIS), the single-outcome PRS approach, and PRSPGx to benchmark their performance against M-Learner in identifying genetic factors explaining HTE. In addition, we applied M-Learner to 45 non-genetic variables measured at baseline (Supplementary Table 7) to see if non-genetic factors can explain any HTE in the same trial.

M-Learner identified significant evidence of HTE (P = 0.001), suggesting that genetic background can modify the response to the bronchodilator therapy (Figure 5A). To ensure robustness, we compared multiple learners—including support vector machine, random forest, elastic net, and neural network—within the M-Learner estimation framework. Support vector machine with a nonlinear radial basis function kernel achieved the best performance, highlighting the importance of employing nonlinear causal machine learning for HTE estimation. In comparison, the SNPxT GWIS, single-outcome PRS, and PRSPGx produced null results (Figure 5A and Supplementary Figure 2). When we applied M-Learner to only non-genetic features, the result was also null (Figure 5A).

**Figure 5:**
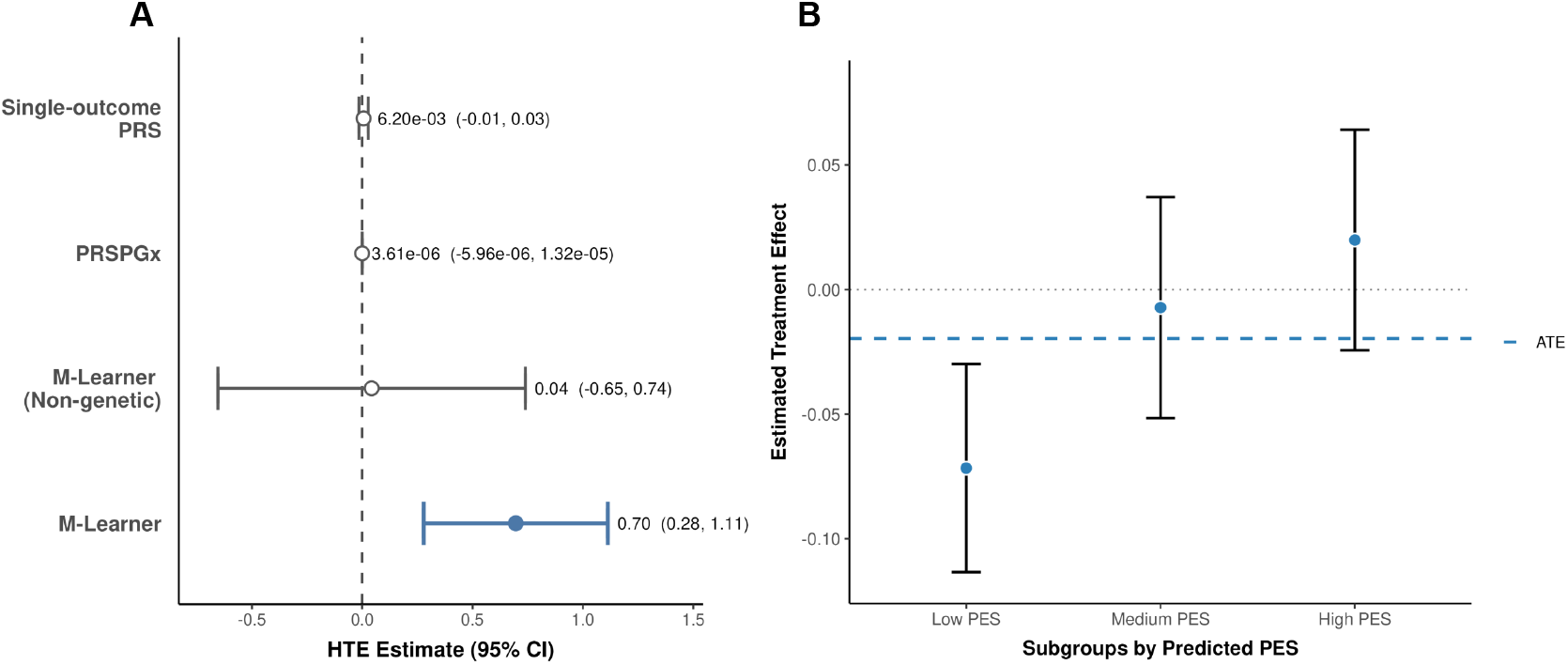
M-Learner identifies HTE of the bronchodilator treatment in smokers. **(A)** M-Learner improves the detection of HTE compared to alternative methods. M-Learner identified a significant HTE (P = 0.001). The single-outcome PRS approach, PRSPGx, and M-Learner using nongenetic variables all failed to identify HTE. **(B)** The average treatment in each subgroup defined by PES strata demonstrate a clear separation: individuals with low PES show negative treatment effects, while those with high PES show a positive response. In particular, individuals in the lowest PES tercile showed a significant but negative treatment effect (P = 0.0008), suggesting potential harm of the bronchodilator treatment in this group.

In addition to detecting overall HTE, we stratified individuals according to their predicted PES and subsequently estimated the average treatment effect within each subgroup defined by the PES tertile (Figure 5B, Supplementary Table 8). The subgroup with the lowest PES exhibited a surprising but statistically significant treatment effect, which led to worse FEV1 (P = 0.0008), suggesting potential harm of the bronchodilator therapy in this group. Although not all subgroups reached statistical significance, the treatment effect estimates in all three strata exhibit a consistent trend, indicating that M-Learner effectively captured the variation in treatment response that was masked in the RCT analysis that only focused on the ATE.

To further interpret the HTE, we examined the PRS that contributed strongly to M-Learner predictions. The PRS for standing height, FEV1, ever smoker, and alcohol use disorder, emerged as the top modifiers of bronchodilator treatment response. These findings indicate that genetic predispositions linked to lung function, substance use, and related physiological traits contribute to the variation in bronchodilator treatment efficacy. These results demonstrate that M-Learner extends beyond global detection of heterogeneity to pinpoint specific genetic mechanisms and pathways that can shape treatment outcomes.

## 3 Discussion

Understanding HTE is crucial for advancing precision medicine, but remains a challenge due to small RCT size, insufficient baseline measures of informative variables, and suboptimal statistical methodology. In this work, we envisioned and demonstrated the potential of using genetic variation to improve HTE inference. We also introduced M-Learner, a causal transfer learning framework that leverages many PRS models pre-trained on external GWAS data to detect and interpret HTE. Through theoretical derivations, simulations, and applications to several RCTs with summary-level or individual-level genetic data, we demonstrated that M-Learner brings important advances to precision therapeutics.

We made two key contributions to the current practice of HTE analysis. First, we argue that future RCTs should routinely incorporate genome-wide genetic variation of trial participants as predictive features, rather than solely relying on non-genetic covariates or a handful of candidate variants—a paradigm we term polygenic efficacy. Our vision is partly based on the fact that the cost for whole-genome genotyping or sequencing continues to drop. But more importantly, genetic information has two unique features that are not shared by other types of data: 1) it is fixed at conception and thus not affected by the treatment; 2) it is known to associate with many human traits, diseases, and behavior. The first feature is important because it enables systematic re-analysis of past RCTs. If genotype data can be generated for trial participants, even if this is done through a new wave of bio-sample collection, genetic variables generated from these samples can be treated as baseline, pre-treatment measures. This will benefit many RCTs, including the ones that have failed in the past due to HTE. If a subgroup of treatment responders can be identified based on genetic information, it could potentially revive failed trials and benefit patients in urgent need for more treatment options. The second feature directly addresses the lack of measured baseline variables that can inform on HTE in RCT data. The idea is that we can use genetic data to “impute” a large collection of human characteristics. Many PRS models have been developed in the literature which can nicely serve this purpose. From a machine learning perspective, this is an excellent example of transfer learning. We can borrow information from the numerous, well-powered GWAS to improve the learning task on HTE. This brings us to the second key contribution we have made in this study. We argue that the estimation of HTE from genetic data should be grounded in a causal transfer learning framework that compresses millions of SNPs into PRS representations, which can then be fine-tuned with trial outcomes to capture the complex genetic modification of treatment response. An important future direction is to explore whether rare genetic variation and genetically predicted transcriptomic, proteomic, and metabolomic variables can further improve HTE inference [38–40].

To operationalize these advances, we developed M-Learner in two forms: M-Learner-I, which uses individual-level genotypes, and M-Learner-S, which relies on summary statistics. M-LearnerI employs sophisticated machine learning to improve the estimation of HTE, even with limited RCT samples. M-Learner-S has broad applications when individual-level genotype data are unavailable from the trial, which is a common struggle in the field. We have demonstrated that through joint consideration of many pre-trained PRS, M-Learner maintains type I error control while achieving markedly higher power compared to other approaches.

Applying M-Learner to RCTs on secukinumab revealed a striking role of BMD genetics in modifying the treatment effect for ankylosing spondylitis. Our results suggest that ankylosing spondylitis patients who are genetically predisposed to lower BMD may receive significantly greater benefit from the anti-IL17 therapy. We believe this is a profound finding for several reasons. First, it provides a clear example where the genetic trait that modifies treatment efficacy is not the genetic predisposition of the primary outcome. We have recently made similar comments on gene-environment interaction research [20], and are now making this observation for HTE inference. Many studies make the (incorrect) assumption that the risk score for the disease being studied is also the modifier of treatment efficacy. Future precision medicine research should expand the search for genetic modifiers into including a broader collection of complex trait candidates. The M-Learner framework provides a principled strategy to implement this idea in real-world RCTs. Second, BMD exhibits a highly specific modifier effect of secukinumab efficacy on ankylosing spondylitis, but not other inflammatory diseases. This observation is consistent with IL17’s central role in osteoimmunology and provides a plausible mechanistic link between inflammatory cytokine signaling and pathological bone remodeling, offering a genetic rationale for prioritizing IL17 blockade in patients with specific skeletal risk profiles. Third, this finding was made using (summary-level) data produced from previous attempts that have generated null results. This highlights the importance of genomic data sharing as well as the need for advanced methods in precision medicine research, especially new strategies that can better handle summary-level genetic association data. Beyond secukinumab, M-Learner also revisited the Lung Health Study, identifying polygenic modifiers of bronchodilator response related to lung function, smoking, and anthropometric traits. Importantly, M-Learner produced PES that can be used to identify responder subgroups for the treatment (in this case, it was a subgroup that showed worse clinical outcomes). These results highlight the potential of using PES to inform trial design, improve sample recruitment, and more broadly, facilitate evidence-based clinical decision making. We believe this is an important step toward a future of healthcare many in the field have envisioned, in which personalized treatment recommendation becomes routine.

Taken together, we present a study that highlights the importance of adopting genetically-driven, hypothesis-free strategies in precision medicine research, and introduce a rigorous statistical framework named M-Learner to facilitate this type of investigation. We expect our findings to motivate important follow-up clinical investigations. We also anticipate M-Learner to have broad applications in genomic medicine and benefit numerous future studies.

## 4 Methods

### Connection between the polygenic GxT model and CATE

We consider the following model for polygenic GxT effects [20]:

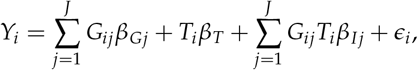

where *Y*_*i*_ is the treatment outcome for individual *i, G*_*ij*_ is the standardized genotype at variant *j* (mean 0, variance 1), 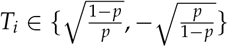 is the standardized treatment indicator with treatment probability *p, β*_*T*_ is the treatment effect, *β*_*Gj*_ and *β*_*Ij*_ denote the additive and interaction effects for variant *j*, and *ϵ*_*i*_ is the error term.

Under this model, the ATE in an RCT can be denoted as

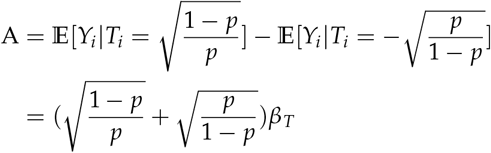

In addition, the CATE can be denoted as

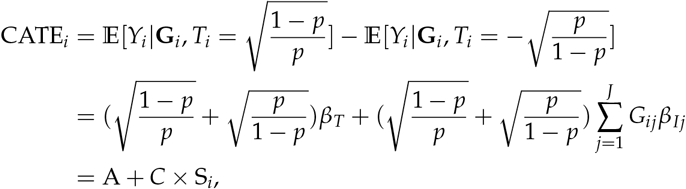

where 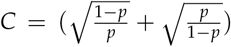 and 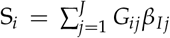. Therefore, the CATE is a linear function of PES. Inferring genetically-driven HTE is equivalent to estimating the PES under the polygenic GxT model.

### M-Learner framework

We introduce a general framework beyond the linear GxT model we have shown above for assessing HTE using causal transfer learning. Let *Y*_*i*_(1) and *Y*_*i*_(0) denote the potential outcomes under treatment and control for *i*-th individual, respectively [41]. Let *T*_*i*_ ∈ {0, 1} represent the binary treatment assignment. The covariate vector contains *J* SNPs denoted as **G**_*i*_. In M-learner, we will compress those *J* SNPs into K pre-trained PRS, 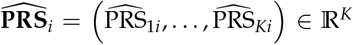, where each 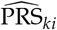 summarizes the polygenic genetic predisposition for a *k*-th trait or disease. PRS can be constructed from external GWAS summary statistics using standard practice [15, 16]. We assume that treatment is randomly assigned conditional on **G**_*i*_, such that the propensity score *e*(**G**_*i*_) = *P*(*T*_*i*_ = 1|**G**_*i*_) is known and bounded away from 0 and 1. In this general setting, CATE for *i*-th individual is defined as

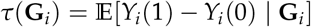

Since individual-level treatment effects are unobservable, we estimate *τ*(**G**_*i*_) using machine learning models combined with causal inference techniques [28].

We adopt a two-stage procedure to estimate CATE. In the first stage, we fit a predictive model using 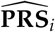 as input to generate individual-level CATE estimates 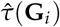. Candidate learners include penalized linear models, support vector machines, random forests, and neural networks, chosen to flexibly capture nonlinear relationships between PRS and treatment response. In the second stage, we evaluate the predictiveness of these estimates using out-of-fold inference. This ensures that each 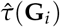 is obtained from a model trained without using the individual’s own outcome data. This cross-fitting strategy mitigates overfitting and yields valid out-of-sample estimates of heterogeneity.

We introduce a pseudo outcome 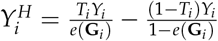. It is well established that 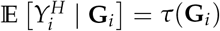 [25, 28], making 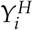 an unbiased proxy for the CATE function. We then estimate predictive models using sequential cross-fitting rather than conventional *K*-fold cross-fitting [27]. Following the sequential validation framework of [42] and [27], the data are partitioned into ordered folds. On fold *k*, we fit the ML model using only the preceding folds 1, …, *k* − 1 and then predict 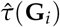 on the current fold *k*. This sequential approach ensures that each prediction is strictly out-of-sample while pooling information across folds with statistical rigor.

To quantify predictiveness and correct for potential bias, we perform a best linear projection of 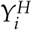 onto 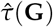

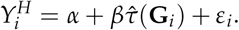

Here, the intercept *α* absorbs the overall ATE component, while the slope *β* captures how much variation in the pseudo outcomes is explained by the predicted CATE. If 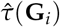 perfectly recovers the true CATE, then *β ≈* 1. If 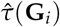 contains only noise, then *β ≈* 0. Rejection of the null hypothesis *β* = 0 provides statistical evidence for HTE explained by genetic variation.

To examine how treatment effects differ across genetically defined risk profiles, we sort and stratify individuals by individuals by their predicted 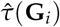 and compute ATEs in each subgroup defined by quantiles of 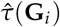.

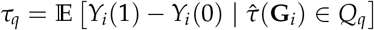

where *Q*_*q*_ denotes the *q*-th quantile bin of estimated 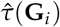. Increasing (or decreasing) *τ*_*q*_ across quantiles indicates a stronger (or weaker) benefit among individuals with higher predicted responses.

For interpretability, we investigate how each individual PRS feature individually relates to HTE. After obtaining out-of-fold 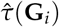, we perform a series of single-variable regressions:

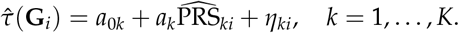

Here, *a*_*k*_ captures the marginal association between 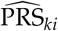 and the predicted treatment effect. To quantify the strength of this association, we define the PRS importance score as the absolute *t*-statistic from the regression:

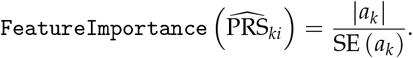

Larger values of `FeatureImportance` 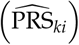 indicate stronger evidence that 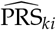 explains

HTE. Ranking PRS by `FeatureImportance` 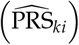 allows us to identify genetic traits most strongly associated with the variation in treatment response.

The M-Learner algorithm proceeds with individual-level data as follows:

#### Algorithm 1: M-Learner-I Algorithm using Individual-level data as input

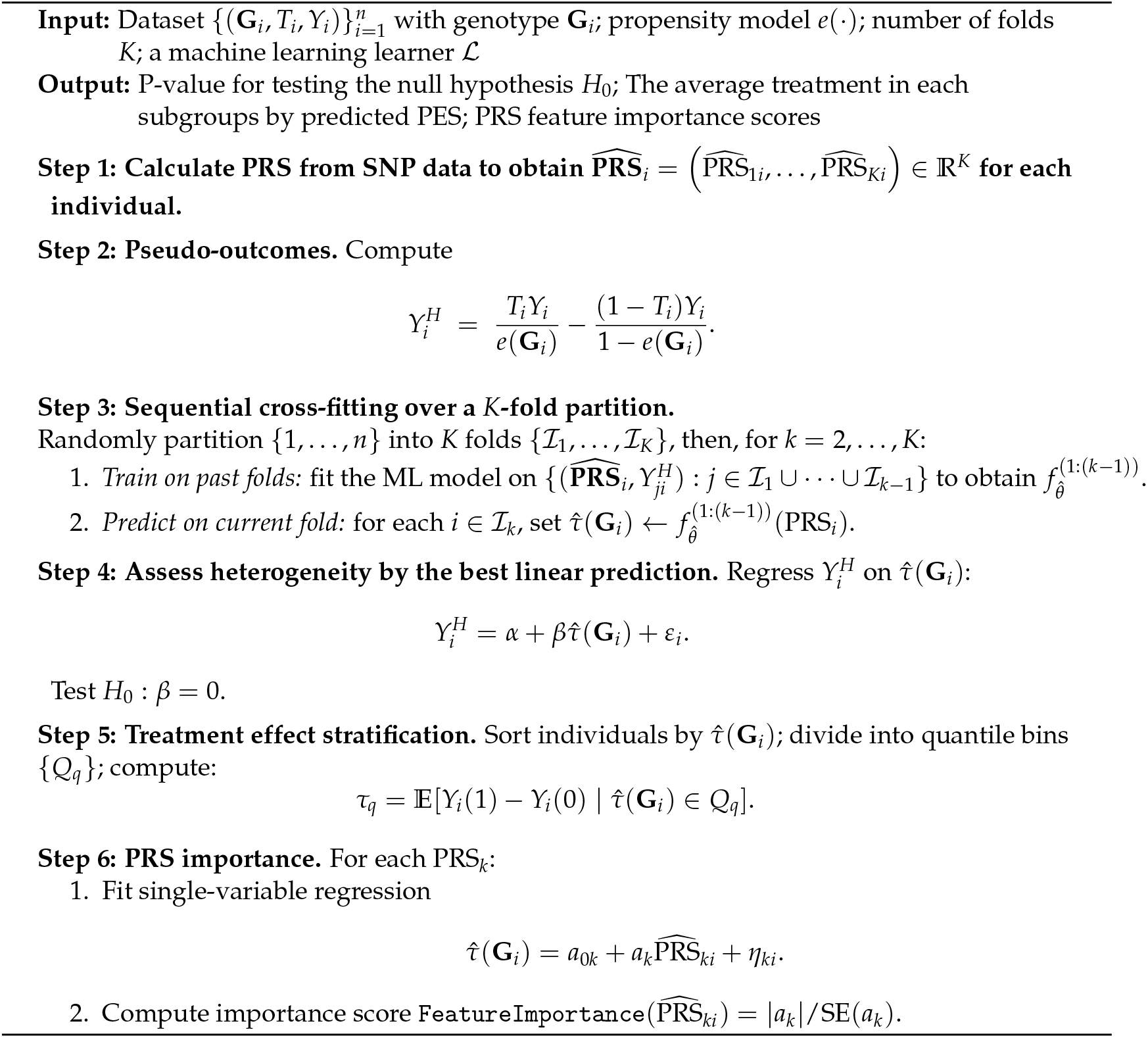

The M-Learner algorithm proceeds with summary statistics as follows:

#### Algorithm 2: M-Learner-S Algorithm using GWIS summary statistics as input

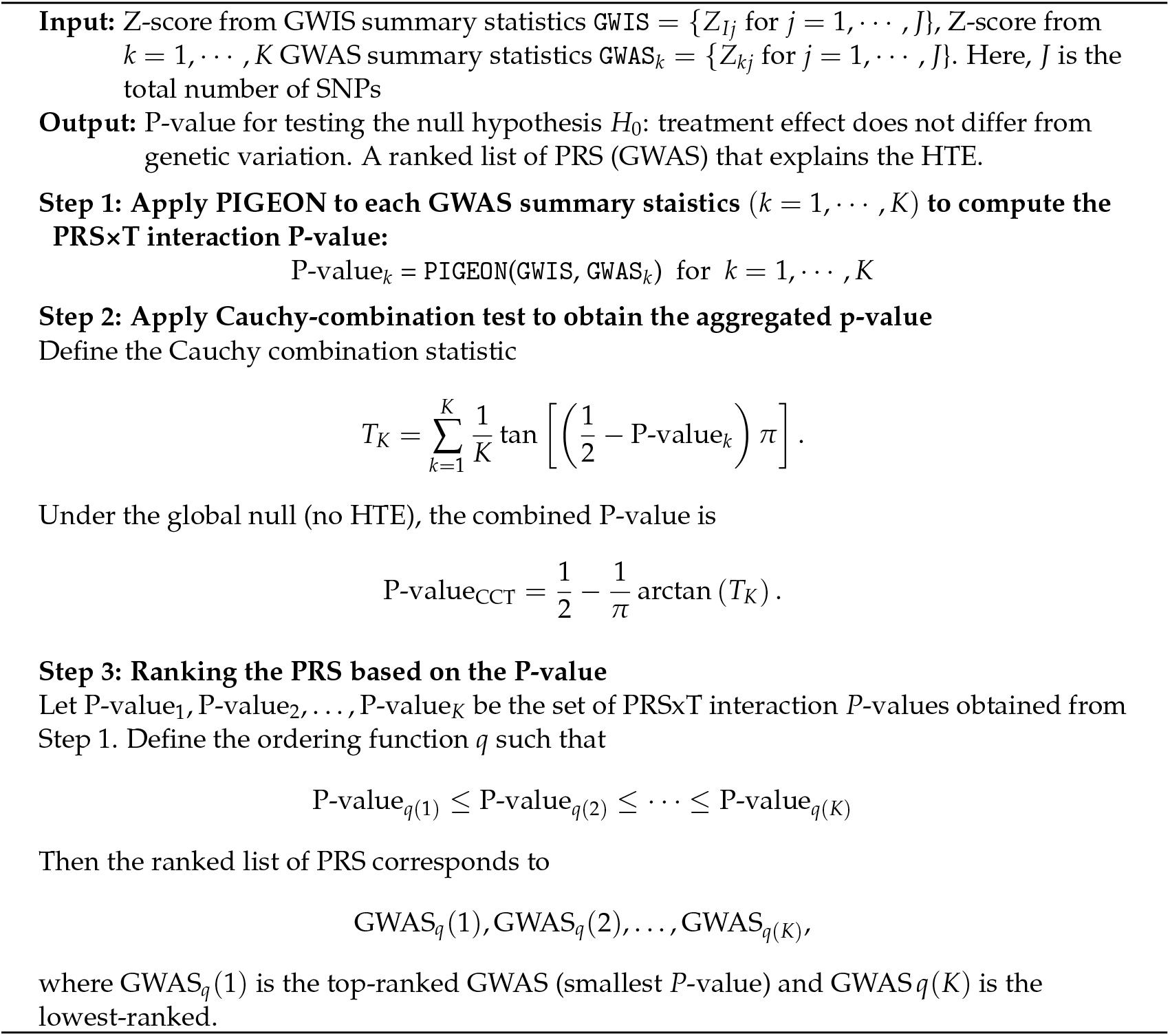

### Derivation for the scaling laws for HTE estimation with genetic variation

We derive the power scaling laws based on the polygenic GxT model. Suppose we have *N*_*test*_ testing samples for quantifying the existence of HTE by linear interaction regression

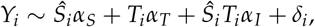

where *Ŝ*_*i*_ is the estimated PES. We use a general notation for *Ŝ*_*i*_ to emphasize that the PES can be estimated in various ways as described below.

We consider different *Ŝ*_*i*_ and evaluate the expected effect size magnitude and statistical power of hypothesis testing of *H*_0_ : *α*_*I*_ = 0 in the test data. We have

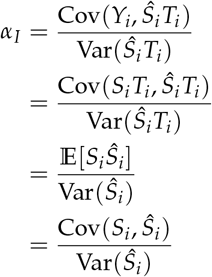

We adopted the random-effects assumption used in the PIGEON framework [20], which is a standard approach in GWAS and G×E studies. The variance-covariance matrix between true PES *S*_*i*_, the estimated PES using within-sample GWIS approach 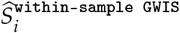 with *N*_*RCT*_ samples, and the estimated PES using a single PRS approach 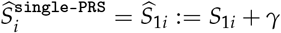 are

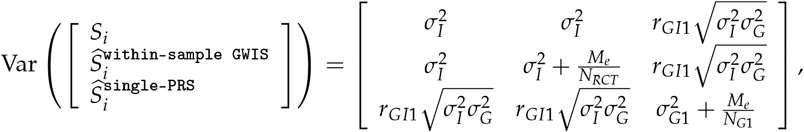

where 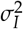 is the proportion of the variance of the treatment response explained by GxT interactions, 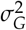 is the heritability for the outcome trait, *r*_*GI*1_ is the correlation between true PES (i.e.,*S*_*i*_) and PRS (i.e.,*PRS*_1*i*_), and *M*_*e*_ is the effective number of SNPs, *γ* is the measurement error in estimating PRS.

Then, if we plug in the estimated PES (i.e. use the SNPs as input and use the estimated interaction coefficient to weight the SNP), we have

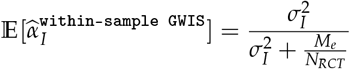

Suppose the *N*_*RCT*_ = 5000, 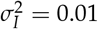, and the typical number for *M*_*e*_ is 60k. If we plug in the estimated PRS, we have

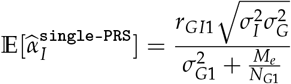

If we have additional K-1 PRS, i.e., 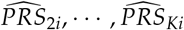, under the same random effect assumption, we have

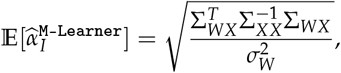

where

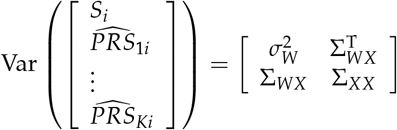

We then have 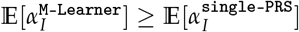.

The statistical power for a two-sided test for *H*_0_ : *α*_*I*_ = 0 can be derived as

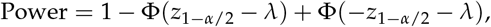

where 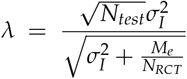 for estimated PES, 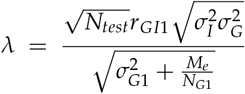 for estimated PRS, and 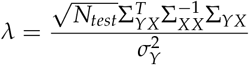 for M-Learner.

### Simulations

Each simulation scenario was repeated 500 times for robust inference. We restricted the analysis to autosomal SNPs that exist in the HapMap3 panel and 1000 Genomes Project. We further filtered SNPs with an imputation quality score greater than 0.9, minor allele frequency (MAF) ≥ 0.05, missing call rate ≤ 0.01, and Hardy–Weinberg equilibrium test p-value ≥ 1.0e-6. We simulated treatment outcome data using 200,000 randomly selected single-nucleotide polymorphisms (SNPs), with effect sizes drawn jointly from a normal distribution. A quantitative phenotype was generated according to

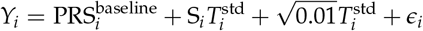

where the binary treatment *T*_*i*_ *∼* Bernoulli(0.5) was standardized to 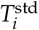 to have a mean of 0 and variance of 1, and *ϵ*_*i*_ denotes Gaussian noise scaled such that Var(*Y*_*i*_) = 1. The term 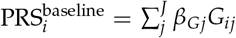 represents the polygenic additive effect that contribute 10% of the treatment outcome, modeled as a single baseline PRS, whereas S_*i*_ specifies the true PES, which can be a linear or nonlinear function of the SNP which we describe below. To simulate realistic genetic correlation structures, we further constructed 25 auxiliary true PRS (i.e., PRS_*ij*_ for *j* = 1, …, 25). Each PRS is simulated with heritability from *N*(0.5, 0.05), and the 25 PRS have correlation coefficients ranging linearly from -0.8 to 0.8. Each PRS is then estimated from a GWAS of 200,000 individuals. The sample size of RCT varies across three scenarios: 500, 1000, and 2000.

1. Linear setting: the true PES was modeled as a weighted sum of the baseline and auxiliary PRS:

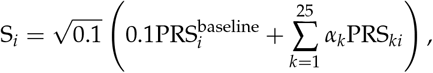

where the weights *α*_*k*_ were equally spaced between -0.6 and 0.6.
2. Nonlinear setting: we replaced the linear combination of auxiliary PRS with a nonlinear transformation:

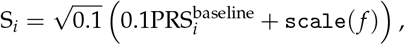

where 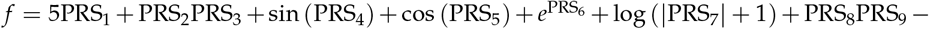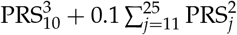, and scale( *f* ) indicates scale *f* to have a mean of 0 and variance of 1. This formulation captures complex, nonlinear SNP-by-treatment interactions that mimic realistic biological heterogeneity.

We evaluated both statistical power (the probability of rejecting the null hypothesis of no HTE at *α* = 0.05) and type I error control under null scenarios with no heterogeneity.

### Analysis of HTE for anti-IL17 therapy in inflammatory diseases

We reanalyzed the SNP-by-treatment GWIS summary statistics derived from clinical and genetic data from 19 RCTs of the anti-IL17 monoclonal antibody secukinumab. The trials covered four indications: psoriasis, psoriatic arthritis, ankylosing spondylitis, and rheumatoid arthritis. After quality control, the dataset included 5,218 individuals (4,063 treated with secukinumab and 1,151 placebo controls).

The primary outcomes were continuous disease activity measures specific to each indication: PASI for psoriasis, DAS28-CRP for psoriatic arthritis and rheumatoid arthritis, and ASDAS-CRP for ankylosing spondylitis. For each participant, we used the change from baseline in outcome score at the primary assessment time point (week 12 for psoriasis; week 16 for psoriatic arthritis, ankylosing spondylitis, and rheumatoid arthritis). Secondary outcomes included laboratory measures, clinician- and patient-reported assessments, and composite indices as described in the original study [10].

For each indication, the original study performed a GxT GWIS using linear regression. Models included treatment (secukinumab vs placebo), genotype dosage, the SNP-by-treatment interaction (the primary term of interest), and covariates (age, sex, BMI, baseline disease activity, three ancestry principal components, and trial-specific design variables). For the present study, we reanalyzed the GWIS summary statistics generated in the original analyses rather than individual-level genotype data. Summary statistics included variant-level regression coefficients, standard errors, and p-values for the GxT interaction terms across all four indications.

### Data analysis in Lung Health Study

We analyzed individual-level clinical and genetic data from the Lung Health Study, which originally enrolled 5,887 smokers aged 35-60 years with early-stage chronic obstructive pulmonary disease [36]. For our analysis, we included 2,441 participants who had both genotype data and complete five-year follow-up of postbronchodilator FEV1. Participants with missing genetic information or incomplete outcome data were excluded, and the separate LHS cohort without smoking intervention was not considered. The primary outcome was defined as the cumulative change in post-bronchodilator FEV1 over five years.

We compared M-Learner to three baseline approaches commonly used for evaluating heterogeneity in treatment response in RCT data. First, a single-outcome PRS approach. We fit a regression model including treatment assignment, the PRS derived from GWAS of FEV1 (the same outcome measured in the trial), and their interaction:

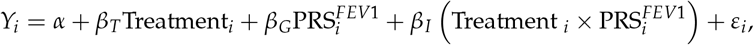

This model tests whether genetic liability to FEV1 modifies treatment response.

We also applied PRSPGx [32], a recently developed framework for pharmacogenomic prediction. At the SNP level, PRSPGx assumes the model

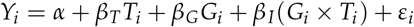

where *G*_*i*_ is genotype for *i*-th individual, *β*_*G*_ and *β*_*I*_ are the main-effect and interaction-effect coefficients estimated from GWIS. PRSPGx then aggregates these SNP-level effects into two polygenic scores: the prognostic score, defined as the weighted sum of genotypes using *β*_*G*_, which captures genetic predisposition to the outcome independent of treatment; and the predictive score, defined as the weighted sum of genotypes using *β*_*I*_, which captures genetic predisposition to differential treatment response. These two scores are conceptually similar to PRS and PES we have discussed throughout the paper. The two scores are then incorporated into the following regression model

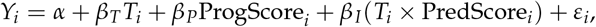

which jointly evaluates the contribution of genetic background to both baseline prognosis and treatment modification. We implemented the GWIS and obtained summary statistics using plink 2.0 [31].

As a baseline, we implemented M-Learner using only non-genetic features. In this setting, the predictors included 45 baseline demographic and clinical variables, such as age, sex, baseline lung function measures, respiratory symptom scores, and other spirometry- and questionnaire-based indicators of respiratory health. These features were combined with treatment assignment to model heterogeneous treatment effects, without incorporating any genetic information.

In addition to these baselines, we applied M-Learner using 4,739 pre-trained PRS from the PGS Catalog [23] as high-dimensional representations of germline genetic variation. These PRS were integrated with randomized treatment assignment and outcome data through sequential crossvalidation to estimate heterogeneous treatment effects and identify the most predictive features. A future direction is to train PRS variables directly from more recent GWAS summary statistics using improved ensemble modeling techniques [43].

## Data Availability

All data produced in the present work are contained in the manuscript

https://qlu-lab.org/data.html

## Code availability

M-Learner is publicly available at https://github.com/qlu-lab/mlearner

## Data availability

The SNP-by-treatment GWIS summary statistics for the anti-IL17 monoclonal antibody secukinumab in inflammatory diseases are available in the GWAS Catalog (https://www.ebi.ac.uk/gwas/, accession numbers GCST90274734 to GCST90274752). Data from the Lung Health study are available by application at https://www.ncbi.nlm.nih.gov/projects/gap/cgi-bin/study.cgi?study_id=phs000291.v2.p1.

## Author contributions

J.Miao., J.Mu., and Q.L. conceived and designed the study.

J.Miao. and J.Mu. developed the statistical framework.

J.Miao., J.Mu., and X.Y. performed the data analysis.

J.F. and L.S. advised on interaction inference and result interpretation.

Q.L. advised on statistical and genetic issues. J.Miao., J.Mu., and Q.L. wrote the manuscript.

All authors contributed to manuscript editing and approved the manuscript.

## Acknowledgments

We acknowledge research support from National Institutes of Health (NIH) grant U01 HG012039, the University of Wisconsin-Madison Office of the Chancellor, and the Vice Chancellor for Research and Graduate Education with funding from the Wisconsin Alumni Research Foundation (WARF).

## Supplementary Information

**Figure S1.**
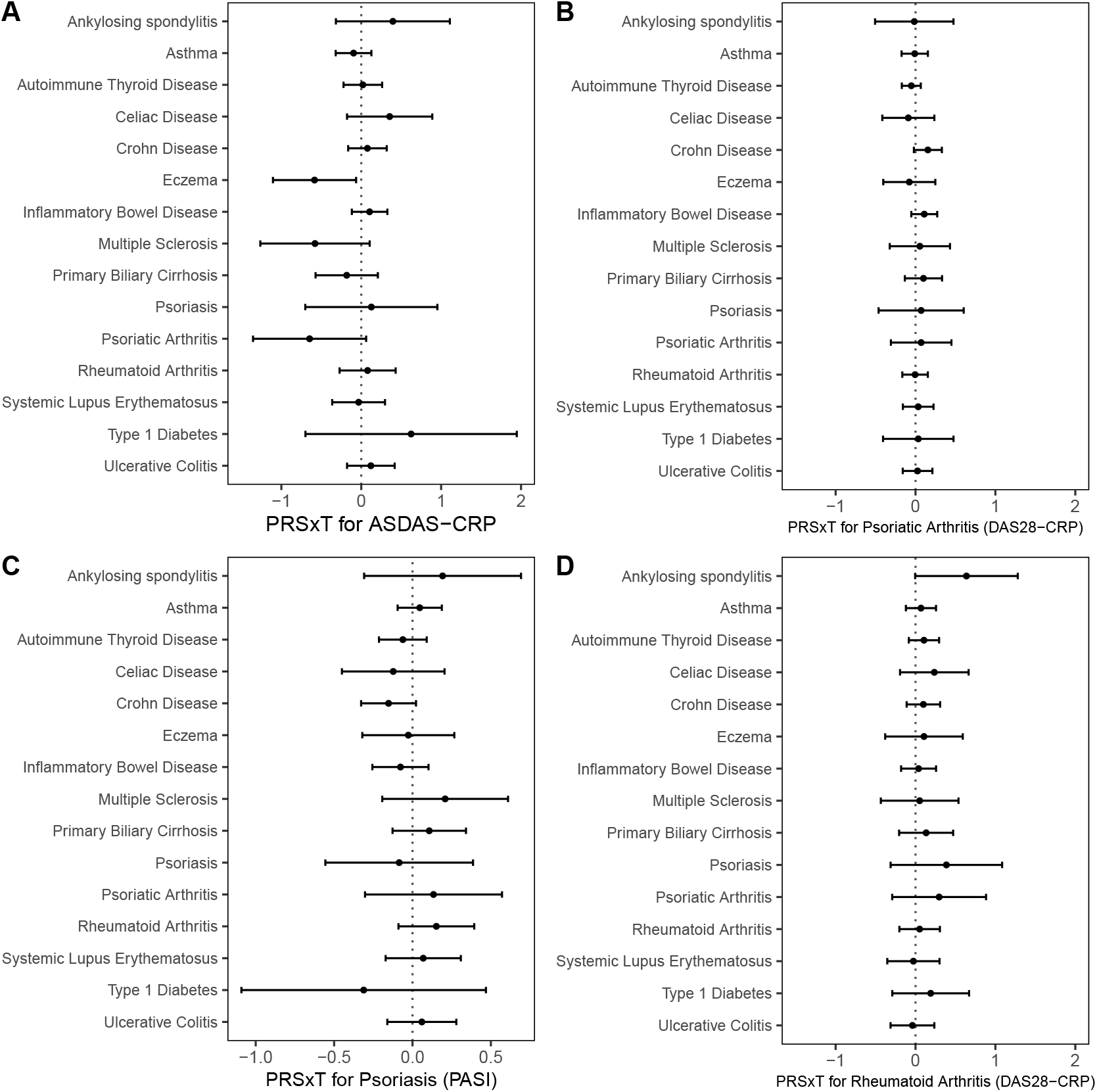
Polygenic risk score by treatment interaction effects (PRS×E) primary outcome for four diseases using PRS for immune-mediated diseases. Points represent estimated interaction effect sizes and horizontal lines denote 95% confidence intervals. The vertical dotted line marks the null (no interaction).

**Figure S2.**
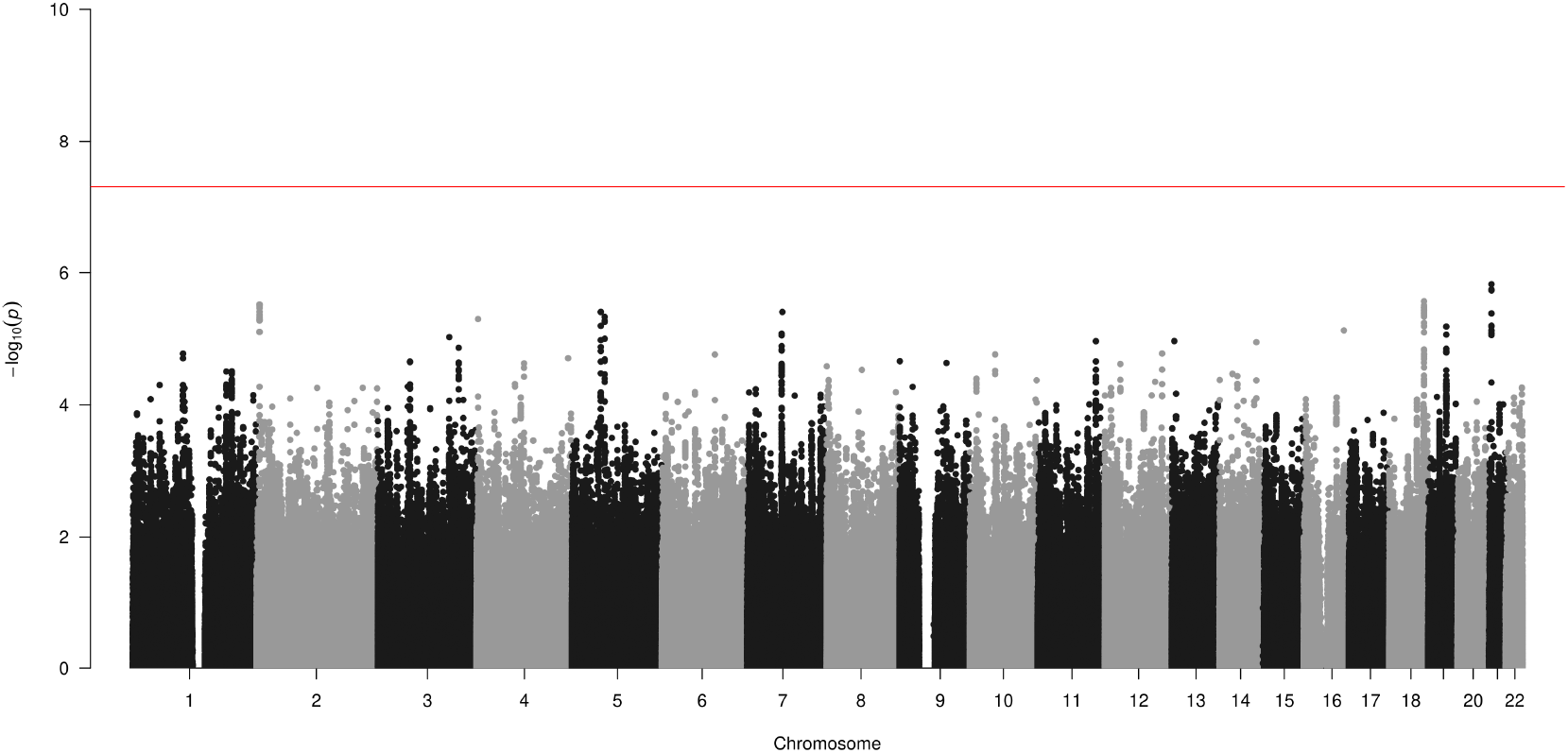
Manhattan plot for genome-wide genotype-by-treatment interaction results in Lung Health Study. The treatment is bronchodilator. The outcome is the changes of FEV1 (Volume that has been exhaled at the end of the first second of forced expiration) from baseline. The red line means P = 5e-8.

